# Issues regarding the Indexing of Adaptive Clinical Trial Articles

**DOI:** 10.1101/2025.03.10.25323694

**Authors:** Neil R. Smalheiser, Andrew Shahidehpour, Ang Michael Troy

## Abstract

Accurate, consistent indexing of biomedical publications is important for retrieving and evaluating evidence, that in turn can affect clinical decisions, evidence syntheses and research directions. Adaptive trials are arguably among the most complex clinical trial designs, being defined as trials in which the allocation or intervention given to subjects is not fixed in advance, but can vary according to the responses or characteristics of the participants. In the course of preparing to tackle the automated indexing of adaptive clinical trial articles, we manually examined the PubMed articles bearing the Adaptive Clinical Trial publication type to date and identified a surprisingly high proportion of erroneous and questionable indexing decisions. If one agrees that each article should be indexed according to its own study design (rather than the study design of the larger trial that it is linked to), then 46.7% of the articles were mis-assigned. If we consider that articles which presented partial results (e.g., one arm of a multi-arm trial) might be broadly deserving of being indexed as adaptive trials, then the adjusted error rate would still be 24.2%. These errors are worth discussing in detail because they have more general lessons to teach us – not only for biomedical end-users who want to utilize indexing terms for efficient retrieval, but for indexers who need to annotate articles accurately and consistently.

## Introduction

Accurate, consistent indexing of biomedical publications is important for retrieving and evaluating evidence, that in turn can affect clinical decisions, evidence syntheses and research directions. Articles are indexed by the National Library of Medicine (NLM) according to one or more Publication Types, e.g., Review, Case Report, or Clinical Trial, and can be described using Medical Subject Headings (MeSH, MH) according to the study design(s) employed in the article, e.g. Case-Control Studies, Random Allocation, or Retrospective Studies (https://www.nlm.nih.gov/mesh/meshhome.html). Our team has carried out machine learning based automated indexing of biomedical articles for more than 50 different publication types and study designs (collectively, PTs) [1] and we are continuing to consider additional PTs to achieve comprehensive indexing. Having created methods for indexing an article as likely to be a Clinical Trial [2], and specifically indexing Randomized Controlled Trials [3, 4], we are now examining methods for indexing articles according to specific subtypes of clinical trials, e.g., phase I, II, III, IV, pragmatic trial, equivalence trial, and adaptive trial.

Adaptive trials are arguably among the most complex clinical trial designs, being defined as trials in which the allocation or intervention given to subjects is not fixed in advance, but can vary according to the responses or characteristics of the participants [5]. Such trials are increasingly common, having been stimulated by the Covid-19 pandemic and the need to be flexible as new possible treatments emerged and as the disease itself evolved in real time [6]. A type of adaptive trial design is the platform trial, in which multiple interventions are all compared against the same control group; as a practical matter, platform trials are considered adaptive, insofar as the list of possible interventions is often flexible, some dropped according to subject responses and new ones added during the trial. NLM newly added Adaptive Clinical Trial as a formal indexing term in 2018, and as of the time of our study, only 182 articles were indexed with that term.

A single clinical trial may give rise to a wide range of multiple publications – some report the overall clinical outcome of the trial, but some articles may only report a small subset of the overall trial. Others may carry out interviews with participants, measure genetic or biochemical markers, re-analyze data across multiple trials, and so on. Trial protocol articles describe the design of a trial in advance. In order to make sense of these diverse papers, two strategies are needed: First, one must have a way to link a given trial to all of its publications, which has motivated several investigators to devise automated linking methods [7-9]. Second, on the other hand, each article should be indexed on its own terms, that is, according to the publication type and study design mentioned in the article itself, regardless of the study design of the underlying clinical trial. This agrees with NLM’s indexing policy for publications.

In the course of preparing to tackle the automated indexing of adaptive clinical trial articles, we manually examined the PubMed articles bearing the Adaptive Clinical Trial publication type to date and identified a surprisingly high proportion of erroneous and questionable indexing decisions, which are worth discussing because they have more general lessons to teach us – not only for biomedical end-users who want to utilize indexing terms for accurate retrieval, but for indexers who need to annotate articles accurately and consistently.

## Methods

Two of us (NRS and AMT) independently annotated all PubMed articles indexed as Adaptive Clinical Trial [Publication Type; PT] as of February 32, 2025. Annotations were resolved to consensus through discussion. Each was scored as either:

A) an Adaptive Clinical Trial article (i.e., presenting trial outcome results that involved adaptive or platform design);
B) a clinical trial article that presented partial results from an adaptive trial, but which did not itself reflect adaptive design (e.g., one arm of a multi-arm study), or that discussed a specific adaptive trial but did not present trial outcome data;
C) a trial protocol or overview of a specific adaptive trial;
D) an article which discussed adaptive trials more generally, e.g., simulated trials, discussions of trial methodology, reviews, or re-analyses of multiple trials;
E) a frank indexing error: articles which were not related to clinical trials at all;
F) a probable indexing error: authors claimed that the article presented results from an adaptive or platform trial, but the available text did not appear to support that claim.

## Results

Out of 182 articles indexed as Adaptive Clinical Trial [PT], only 97 were actually scored as being an adaptive clinical trial. 41 were presenting partial results or discussing specific trials, 14 were protocol or overview articles, 11 were more general discussions of adaptive trials, 14 were frank errors, and 5 were probable errors. If one agrees that each article should be indexed according to its own study design (rather than the study design of the larger trial that it is linked to), then (182-97)/182 = **46.7%** of the articles were mis-assigned! This is a surprisingly high error rate. If we consider that the 41 articles which presented partial results might be broadly deserving of being indexed as adaptive trials, then the adjusted error rate would be reduced to (182-97-41)/182 = **24.2%**, still very high.

Many of the mis-assigned articles should have properly been indexed as “Clinical Trials as Topic”[MH] or “Clinical Studies as Topic”[MH], reflecting the fact that they are discussing trials rather than presenting clinical outcomes. This includes trial protocols, for whom NLM indexing policy would assign “Clinical Trial Protocol”[PT], the vast majority of which are also indexed as “Clinical Trials as Topic”[MH] and “Clinical Studies as Topic”[MH]. We found that a total of (14+11) = 25 articles should have been indexed “as topic” instead, whereas only 6 of the 182 articles were actually dual indexed as “Clinical Trials as Topic”[MH] or “Clinical Studies as Topic”[MH], and none at all were dual indexed as “Adaptive Clinical Trials as Topic”[MH].

The frank errors were disproportionately indexed in the first few years after introduction of the indexing term (e.g. 9 of the 14 were published in or before 2019) which may indicate some uncertainty as the indexing process was developed. However, as shown in Table 1, these articles include surveys given to faculty, genetic sequencing studies, and even materials science! It is hard to imagine how such a crazy mix of articles received assignments as Adaptive Clinical Trial.

**Table 1.**
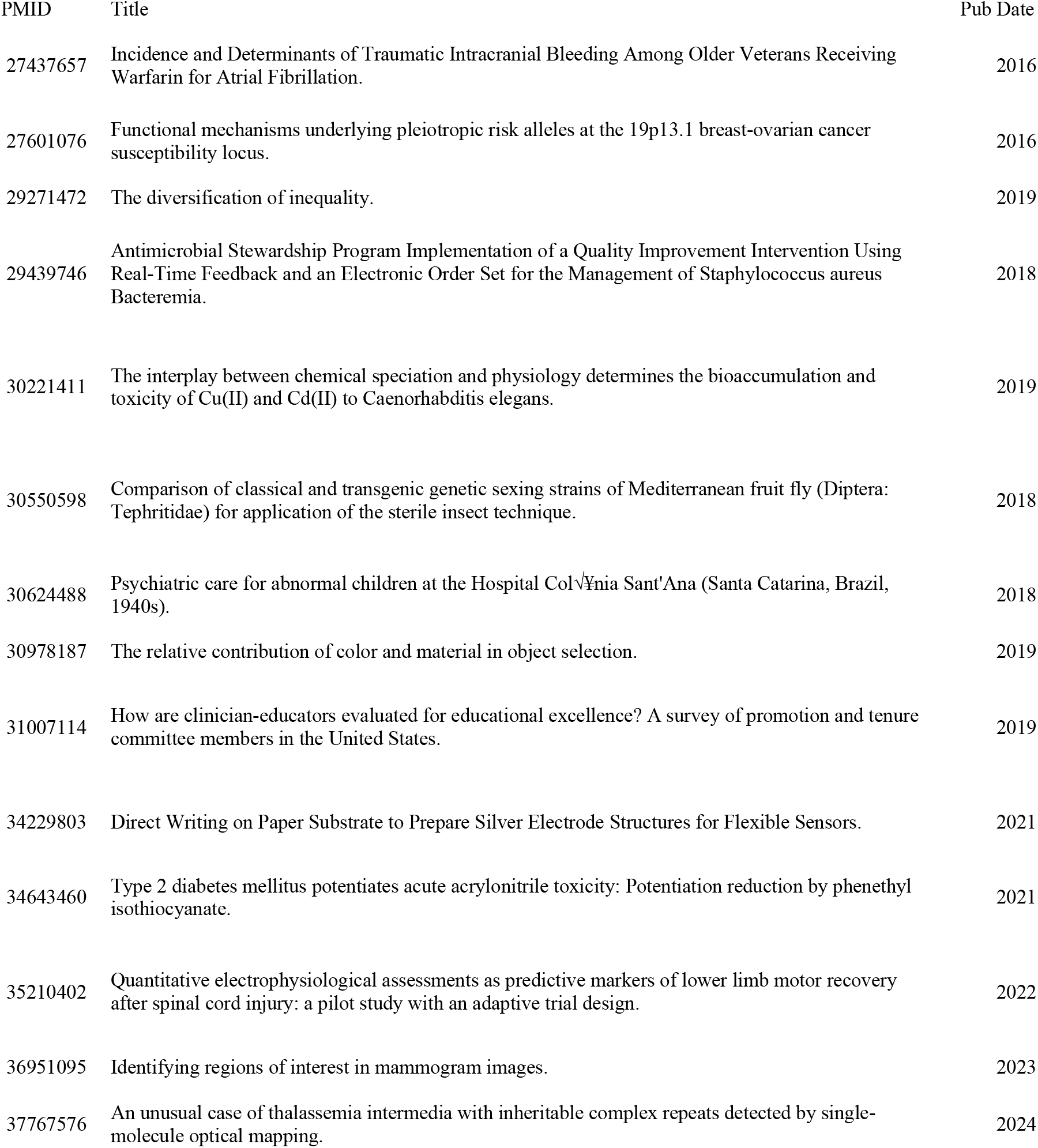
Frank errors in indexing of Adaptive Clinical Trial articles.

These are errors of commission. What about errors of omission, when articles that deserve to be indexed as Adaptive Clinical Trial fail to receive that assignment? Given that the indexing term was only introduced in 2018, the fact that earlier articles may not have been indexed as such is not necessarily an oversight nor an error. However, to estimate informally how many adaptive clinical trial articles may be in PubMed overall, we carried out a PubMed search to identify any PubMed articles indexed as Clinical Trial[PT] which mentioned either “adaptive trial” OR “platform trial” and were not already indexed as “Adaptive Clinical Trial”[PT]. Only 34 articles were retrieved, the majority published before 2018, suggesting there are not a lot of relevant articles missed in the current indexing effort. Retrospective indexing of publications, as carried out by NLM in many cases, may hopefully assign the earlier articles in due course.

As an update, before submitting this paper, we double-checked the latest status of indexing (as of March 9, 2025) and found that three of the articles that we had judged “as topic” had their indexing changed -- removing Adaptive Clinical Trial[PT] and adding “as topic” MeSH terms. This indicates that NLM is carrying out at least a limited extent of error correction on articles.

## Discussion

Adaptive clinical trials have complex designs, and the publications that are linked to them appear to represent a challenge for indexers, since our analyses show a surprisingly high error rate for NLM indexing of these articles. Depending on whether one makes a strict or narrow interpretation of indexing policy, about a quarter to one-half (24.2% to 46.7%) of assignments were inappropriately made. This is in contrast to our earlier estimates of NLM indexing accuracy of randomized controlled trials (RCTs), in which only about 5% of articles marked as RCTs were judged not to be indexed appropriately, and another 3% additional RCT articles failed to receive RCT indexing [3].

The largest category of indexing errors involved articles which discussed trials (e.g., describing trial designs, proposing trial design methodology, or reviewing multiple trials) but did not present the clinical outcomes of a specific adaptive clinical trial. These articles would properly be indexed as “Adaptive Clinical Trials as Topic”[MH]. An additional category were frank errors (Table 1) for which no obvious explanation is forthcoming. There are several lessons to be learned here:

First, for biomedical end-users seeking to retrieve the full range of published articles related to a specific adaptive clinical trial, or related to this class of trials in general: Is it really so bad to include extra articles in a PubMed search, as long as most of the appropriate articles are retrieved? Perhaps not when the class includes only 182 articles, but indexing schemes must apply consistently to all article types, which may include 1,000, 5,000 or 50,000 publications.

Better that indexing is not overly inclusive. However, it is indeed important to be able to identify the entire assemblage of articles that are generated by, or related to, a given trial. For that purpose, it would suffice for authors to mention the trial registry number in each publication. As well, one can employ linking tools such as Trials to Publications [9] which attempt to identify publications linked to registered trials even when registry trial numbers are not explicitly given, or missing from the abstract and only given within the full-text of the article [10].

Second, for those of us interested in automated indexing of adaptive trials, one must be careful to remove the “as topic” articles and the frank errors from the overall set of PubMed indexed articles, before using them as a training set for “Adaptive Clinical Trial”. On the other hand, we note that there are currently 552 PubMed articles indexed as “Adaptive Clinical Trials as Topic”[MH], several times more than are indexed as Adaptive Clinical Trial [PT]. This suggests that is may be worthwhile to carry out automated indexing for “Adaptive Clinical Trials as Topic” as well.

Finally, a limitation of this study is that the indexing criteria and notes given to NLM indexers are not public, so at present we are left to infer these from the observed indexing behavior. Definitions for publication types and study designs are given in the MeSH database (https://www.nlm.nih.gov/mesh/meshhome.html) but these are clearly not detailed enough for indexers. For those who maintain bibliographic databases and annotate articles with indexing terms and other metadata, permitting and incorporating public feedback regarding detection of errors would improve the accuracy of indexing. Perhaps current bibliographic databases resemble static data resources, but in the future, they could instead interface dynamically with the users and become interactive.

## Data Availability

All data produced are available online at PubMed.gov

## Abbreviations

NLM: National Library of Medicine.
MH: Medical Subject Heading.
MeSH: Medical Subject Heading.
PT: publication type.

## References

1. Cohen AM, Schneider J, Fu Y, McDonagh MS, Das P, Holt AW, Smalheiser NR. Fifty ways to tag your PubTypes: Multi-tagger, a set of probabilistic publication type and study design taggers to support biomedical indexing and evidence-based medicine. medRxiv. 2021 Jul 16:2021–07.

2. Menke JD, Kilicoglu H, Smalheiser NR. Publication Type Tagging using Transformer Models and Multi-Label Classification. medRxiv 2025; doi: 10.1101/2025.03.06.25323516.

3. Cohen AM, Smalheiser NR, McDonagh MS, Yu C, Adams CE, Davis JM, Yu PS. Automated confidence ranked classification of randomized controlled trial articles: an aid to evidence-based medicine. J Am Med Inform Assoc. 2015 May;22(3):707–17. doi: 10.1093/jamia/ocu025.

4. Wallace BC, Noel-Storr A, Marshall IJ, Cohen AM, Smalheiser NR, Thomas J. Identifying reports of randomized controlled trials (RCTs) via a hybrid machine learning and crowdsourcing approach. J Am Med Inform Assoc. 2017 Nov 1;24(6):1165–1168. doi: 10.1093/jamia/ocx053.

5. Ben-Eltriki M, Rafiq A, Paul A, Prabhu D, Afolabi MOS, Baslhaw R, Neilson CJ, Driedger M, Mahmud SM, Lacaze-Masmonteil T, Marlin S, Offringa M, Butcher N, Heath A, Kelly LE. Adaptive designs in clinical trials: a systematic review-part I. BMC Med Res Methodol. 2024 Oct 4;24(1):229. doi: 10.1186/s12874-024-02272-9.

6. Glasziou P, Sanders S, Byambasuren O, Thomas R, Hoffmann T, Greenwood H, van der Merwe M, Clark J. Clinical trials and their impact on policy during COVID-19: a review. Wellcome Open Res. 2024 Jan 30;9:20. doi: 10.12688/wellcomeopenres.19305.1.

7. Goodwin TR, Skinner MA, Harabagiu SM. Automatically linking registered clinical trials to their published results with deep highway networks. AMIA Jt Summits Transl Sci Proc 2018; 2017: 54–63.

8. Dunn AG, Coiera E, Bourgeois FT. Unreported links between trial registrations and published articles were identified using document similarity measures in a cross-sectional analysis of ClinicalTrials.gov. J Clin Epidemiol 2018; 95: 94–101.

9. Smalheiser NR, Holt AW. A web-based tool for automatically linking clinical trials to their publications. J Am Med Inform Assoc. 2022 Apr 13;29(5):822–830. doi: 10.1093/jamia/ocab290.

10. Holt AW, Troy AM, Smalheiser NR. Distribution of trial registry numbers within full-text of PubMed Central articles: implications for linking trials to publications and indexing trial publication types. Trials. 2025 Jan 31;26(1):34. doi: 10.1186/s13063-025-08741-w.

